# Investigating the attitudes of Canadian paramedic students towards homelessness

**DOI:** 10.1101/19003483

**Authors:** Alyson Cochrane, Priya Pithia, Emma Laird, Kelly Mifflin, Venessa Sonley-Long, Alan M. Batt

## Abstract

**Introduction:** When paramedics are dispatched, it is expected that every patient receives the same level of care regardless of variable factors. Homelessness is a growing social issue across Canada that is particularly prevalent in urban areas. The quality of healthcare delivered to individuals experiencing homelessness may be influenced by negative attitudes held by healthcare professionals. There is an absence of literature quantifying the perspectives of paramedics towards homelessness; therefore, the focus of this study was to identify the attitudes of paramedic students towards homelessness and to continue the conversation in regards to the evolving educational needs of paramedic students.

**Methods:** This study employed a longitudinal design of a convenience sample of first year paramedic students in a college program in Ontario, Canada. The ‘Health Professional’s Attitude Towards the Homeless Inventory’ (HPATHI) was distributed to participants before and after placement and clinical exposure. The questionnaire includes 19 statements which participants respond to on a Likert scale. Mean scores were calculated, and statements were categorized into attitudes, interest, and confidence. Data were collected post-placement on interactions with persons experiencing homelessness.

**Results:** A total of 52 first year paramedic students completed the HPATHI pre-placement and 47 completed the questionnaire post-placement. Mean scores for attitudes (pre 3.64, SD 0.49; post 3.85, SD 0.38, p=0.032), interest (pre 3.91, SD 0.40; post 3.84, SD 0.39,p=0.51) and confidence (pre 4.02, SD 0.50; post 3.71, SD 0.67, p=0.004) were largely positive, but there was a demonstrated decreasing trend in confidence with, and interest in, working with those experiencing homelessness. Participants reported an average of 60 hours of placement, during which 15 participants (32%) reported interactions with people experiencing homelessness.

**Conclusion:** First year paramedic students demonstrate overall positive attitudes towards those experiencing homelessness, and the mean score for attitudes improved over the surveys. However, there were demonstrable decreases in confidence and interest over time, which may be related to the type and frequency of interactions during clinical placement. Paramedic education programs may benefit from the inclusion of focused education on homelessness, specific clinical experiences, and education related to social determinants of health.

## Introduction

Homelessness is a pervasive social issue across Canada. It is particularly visible in urban areas, and is a growing issue in many western countries. Over 235,000 Canadians experience homelessness every year, while an estimated 150,000 access emergency shelters every year. At least 35,000 Canadians are homeless on any given night, while approximately 50,000 are part of the “hidden homeless” -provisionally accommodated by friends or relatives as they have nowhere else to go (Gaetz et al. 2016). 9,200 people are homeless in Toronto on a given night (City of Toronto 2018), with 7,530 in Ottawa, 1,519 in Winnipeg, 2,911 in Calgary (Canadian Observatory on Homelessness 2019a), and recent figures identified over 2,200 are homeless on any given night in Vancouver (Marshall 2019). This is not a problem that is restricted to large urban centres however - smaller population centres, remote, and Northern communities across Canada also report increasing levels of homelessness (Canadian Observatory on Homelessness 2019a), and homelessness in these areas comes with its own unique challenges that are poorly addressed in the literature.

Historically, individuals experiencing homelessness in Canada were older, single men. However, homelessness today is much more diverse. Women, families and youth are experiencing homelessness more than in the past (Gaetz et al. 2016). Almost 40,000 youth experience homelessness in Canada, and between 25-40% of this population identify as LGBTQS (Abramovich and Shelton 2017; The 519 2019). A significant proportion of homeless persons are Indigenous (∼16-24%), and the number of older adults and seniors experiencing homeless is also growing (∼25%) (Gaetz et al. 2016). These changing demographic profiles also present new challenges when considering social services and healthcare access for this population. For example, 1 in 3 transgender individuals are rejected from shelters due to their gender identity or gender expression (Canadian Observatory on Homelessness 2019b), while older adults who are experiencing homelessness are more likely to have poorer health, and mental health issues (Kimbler et al. 2017).

Living homeless affects many individuals’ overall health and quality of life, including their mental health and their ability to adequately manage chronic conditions (Frankish et al. 2005). Those experiencing homelessness have high rates of physical and mental illness, and they have increased rates of morbidity and mortality when compared to the general population (Frankish et al. 2005; Hwang and Bugeja 2000). Due to the difficulty in accessing general healthcare and social services, many persons experiencing homelessness regularly interact with paramedic services and emergency departments (ED) as their primary source of healthcare (Goering et al. 2002; Moore et al. 2011; Padgett et al. 1995; Pearson et al. 2007). Approximately 50% of those experiencing homelessness are affected by a wide range of chronic medical issues, some of which include diabetes, chronic obstructive pulmonary disease, arthritis, musculoskeletal disorders, and skin and foot problems (City of Toronto 2018). Management of such chronic medical issues while homeless is challenging (Hwang and Bugeja 2000), and results in repeated access to healthcare services through emergency access points such as paramedic services and EDs for chronic issues. This utilization of healthcare services may be viewed as “inappropriate” and may subsequently lead to provider prejudice. When those experiencing homelessness seek healthcare in such a manner, they often report feelings of being “unwelcome” by healthcare workers (Wen et al. 2007). They describe their experiences as dehumanizing and disempowering.

Existing literature suggests that healthcare professionals demonstrate overall negative attitudes towards those experiencing homelessness and towards individuals of lower socioeconomic status (Fine et al. 2013; Minick et al. 1998; Price et al. 1989; Sibley et al. 2017; Zrinyi and Balogh 2004). Negative experiences with paramedics described by homeless shelter users were predominantly related to provider attitude (Leggio et al. 2019). These experiences of prejudice and negative attitudes may act as a barrier to accessing healthcare in the future (Lester and Bradley 2001; Parkinson 2009; Wen et al. 2007). By contrast, when staff have a welcoming attitude, persons experiencing homelessness feel valued and part of society (Wen et al. 2007). Positive experiences are not the only benefit of compassionate care. In fact, when those experiencing homelessness in Toronto experienced compassionate care in the ED, it resulted in a significant decrease in both the total number of return visits by 28% (95% CI 14-40%, p=0.001), and the average frequency of visits per month by 33% (95% CI 29-44%, p<0.001) (Redelmeier et al. 1995). This result reflects an economic benefit for the healthcare industry as well as a positive influence on patient care.

There is a distinct lack of literature that explores the intersection of paramedicine and homelessness. In particular, the attitudes of paramedics towards persons experiencing homelessness, their knowledge and understanding of health and social issues related to homelessness, and the role that paramedics may be able to play in providing healthcare for those experiencing homelessness remain poorly investigated and reported. Those experiencing homelessness have previously indicated their trust in paramedics (Zakrison et al. 2004), and anecdotally paramedics interact regularly with individuals experiencing homelessness. Paramedics are therefore uniquely positioned to care for this population – they are afforded the ability to assess the health and social needs of patients in context, and can refer such individuals to a variety of alternative services and destinations outside of the ED as appropriate (Moore et al. 2011; Tangherlini et al. 2016). Given the diversity of values, beliefs and perspectives on homelessness (Frankish et al. 2005), it is important to identify if negative attitudes exist among paramedics, as quality of care may otherwise be influenced. Therefore, the purpose of this study is to investigate the attitudes of paramedic students towards homelessness and to discuss potential opportunities within paramedic education to improve healthcare delivery for those experiencing homelessness.

## Methods

### Terminology

We appreciate that there are differing views on the terminology used in relation to homelessness. Throughout this manuscript we have elected to use the term “experiencing homelessness”, as there are varying definitions of homelessness, and it can be chronic or transient in nature (Rich 2017). Homelessness also affects different groups of people in different ways, and every individual’s experience is unique (Canadian Observatory on Homelessness 2019c).

### Participants

This study was a longitudinal study of a convenience sample of first year paramedic students in a two-year paramedic diploma program at [redacted for peer review] in Ontario, Canada. All students were invited to participate in the study. Participants were provided with an explanatory statement prior to participation, and were informed that participation was entirely voluntary and responses were completely anonymous.

### Ethical considerations

Ethics approval was granted by the Research Ethics Board at [redacted for peer-review] (approval S 18-06-28-1).

### Materials

This study utilized a paper-based questionnaire, the Health Professional’s Attitude Towards the Homeless Inventory (HPATHI) (Buck et al. 2005), a 19-statement survey which participants respond to on a 5-point Likert scale ranging from strongly disagree (1) to strongly agree (5). The HPATHI instrument is a validated tool that has previously been used in other healthcare professions (Crow 2014). It has not previously been tested in paramedic students. We elected to use the HPATHI to allow for comparisons with existing literature in other professions, and due to the ability to group responses into attitudes, interest, and confidence (Fine et al. 2013).

### Data collection

The questionnaire was distributed to participants on two occasions - the beginning of the first semester before clinical placement experience, and at the beginning of the second semester after clinical placement experience. No demographic data were collected. Responses were entered into SPPS v.20 (IBM Corp., 2011) for collation and analysis.

### Data analysis

Negatively worded statements in the HPATHI (statements 4, 5, 10 and 16) were reverse-scored so that for every survey item, a higher numerical score (towards 5) indicated a more positive attitude towards homelessness. Statement 19 is excluded from analysis due to irrelevance to paramedicine. Descriptive statistics (mean, SD) are used to describe trends in the data and to discuss our results in the context of the existing literature, and t-tests were used to compare means before and after clinical placement. All analyses were 2-tailed and p<0.05 was considered significant.

### Clinical placement setting

The majority of these students completed clinical placement experience in the area surrounding the city of London. This city identified approximately 400 people experiencing homelessness, with 30% identified as Indigenous (Canadian Observatory on Homelessness 2019a). 60% of those surveyed experienced chronic homelessness *(experienced homelessness for six months or more in the past year)*. The majority of respondents listed abuse or relationship issues as the cause for their homelessness (City of London 2017a). A total of 43% reported chronic health issues, and 56% reported emergency shelters as the place they slept most frequently. In the past five years, 10,782 people have stayed at homeless shelters in this city. Of these, 66% were adult males, 21% were adult females, 11% were dependent children and youth, and 2% did not report their gender. Individuals staying at shelters ranged in age from 15 to 96, with an average age of 39. Young adult females (≤29 years of age) comprised 42% of females accessing shelters. The average number of nights stayed increased from 24 in 2011 to 41 in 2016, with males on average staying longer than females (City of London 2017b). These statistics are reflective of the urban city of London and do not account for those experiencing homelessness in surrounding rural centres.

## Results

A total of 52 first year paramedic students completed the HPATHI pre-placement and 47 completed the questionnaire post-placement. Table 1 outlines the mean scores (pre- and post-placement) of each statement from the HPATHI questionnaire (excluding question 19).

**Table 1.** Mean scores (pre- and post-placement) of each HPATHI statement.

| No. | Pre-placement mean (SD) | Post-placement mean (SD) | p value | No. | Pre-placement mean (SD) | Post-placement mean (SD) | p value |
| --- | --- | --- | --- | --- | --- | --- | --- |
| 1 | 3.51 (0.86) | 3.72 (0.58) | 0.14 | 10 | 3.35 (0.84) | 2.87 (0.90) | 0.008 |
| 2 | 4.49 (0.58) | 4.66 (0.48) | 0.11 | 11 | 3.25 (0.82) | 3.47 (0.83) | 0.20 |
| 3 | 4.24 (0.76) | 4.21 (0.69) | 0.87 | 12 | 3.45 (0.99) | 3.89 (0.87) | 0.02 |
| 4 | 3.47 (1.19) | 3.77 (1.11) | 0.20 | 13 | 4.59 (0.64) | 4.47 (0.65) | 0.36 |
| 5 | 3.71 (0.64) | 3.77 (0.96) | 0.71 | 14 | 3.84 (0.81) | 3.81 (0.82) | 0.82 |
| 6 | 2.88 (0.99) | 3.38 (0.85) | 0.008 | 15 | 3.51 (0.92) | 3.40 (0.90) | 0.56 |
| 7 | 3.94 (0.95) | 3.51 (1.06) | 0.03 | 16 | 3.76 (0.74) | 3.72 (0.90) | 0.80 |
| 8 | 4.35 (0.63) | 4.38 (0.53) | 0.79 | 17 | 3.84 (0.70) | 3.81 (0.88) | 0.83 |
| 9 | 4.45 (0.54) | 4.09 (0.83) | 0.01 | 18 | 3.78 (0.97) | 3.81 (0.82) | 0.89 |

These statements were then grouped into three different categories outlined in the existing literature: attitudes (statements 1-6, 11, 12, 18), interest (13-17), and confidence (7-10). Mean scores for attitudes, interest and confidence were largely positive, but there was a demonstrable decreasing trend across the surveys related to interest and confidence in working with people experiencing homelessness. Table 2 outlines the mean scores (pre- and post-placement) for each category.

**Table 2.** Mean scores (pre- and post-placement) for each category.

| Attitudes (1-6, 11, 12, 18) |  | Interest (13-17) |  | Confidence (7-10) |  |
| --- | --- | --- | --- | --- | --- |
| Pre-placement mean (SD) | Post-placement mean (SD) | Pre-placement mean (SD) | Post-placement mean (SD) | Pre-placement mean (SD) | Post-placement mean (SD) |
| 3.64 (0.49) | 3.85 (0.38) | 3.91 (0.40) | 3.84 (0.39) | 4.02 (0.50) | 3.71 (0.67) |
| $p=0.032$ | | $p=0.51$ | | $p=0.004$ | |

Participants reported an average of 60 hours of placement in the post-placement survey. Participants were also surveyed on whether they interacted with people experiencing homelessness during their placement (Table 3).

**Table 3.**
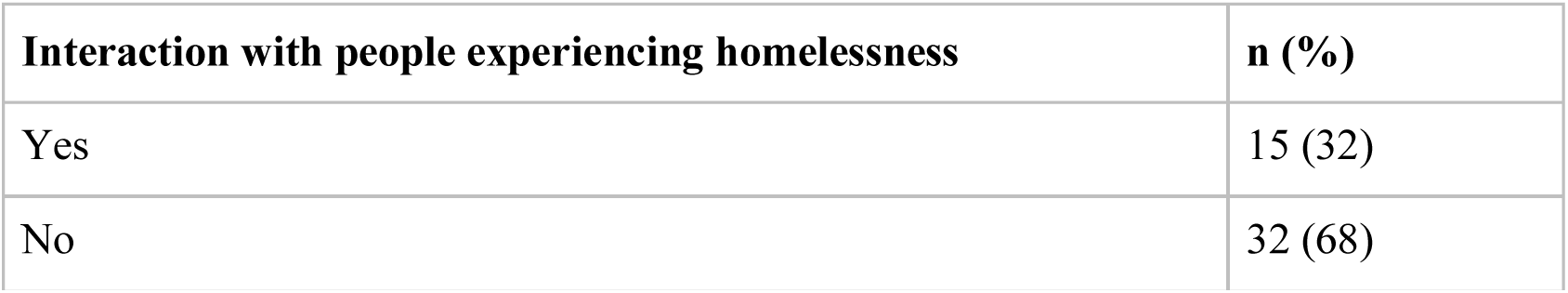
Self-reported interaction with people experiencing homelessness during placement.

| Interaction with people experiencing homelessness | n (%) |
| --- | --- |
| Yes | 15 (32) |
| No | 32 (68) |

## Discussion

Our study demonstrated that first year paramedic students had overall positive attitudes towards people experiencing homelessness, and the mean score for attitudes improved over the surveys (p=0.032, see Table 2). However, there appeared to be a decreasing trend over time in relation to confidence in working with (p=0.004), and interest in working with (p=0.51) those experiencing homelessness (Table 2). Of note within the individual statements was a statistically significant improvement in participants’ perceptions of the healthcare needs of the underserved (statements 6 and 12, see Table 1). However, comfort levels (and thereby confidence) demonstrated a statistically significant decline (statements 7, 9 and 10, see Table 1). In addition, only 32% (n=15) of participants directly interacted with people experiencing homelessness during their placement (Table 3).

Professional, non-judgmental, compassionate care toward homeless shelter users appeared to best describe the difference between a positive experience from a negative one for the individual when interacting with paramedics (Leggio et al. 2019). The positive attitude towards those experiencing homelessness demonstrated by the participants in our study is encouraging, and provides a foundation for continued compassionate care. The decline in interest and confidence in working with people experiencing homelessness may have been related to the type and frequency of interactions during clinical placement. One reason for this decreasing trend could be a lack of formal education surrounding the health and social needs of those experiencing homelessness. Indeed, social determinants of health in general are poorly addressed in the curriculum at present. Given the growing recognition that social and economic factors shape individuals’ health outcomes, it seems prudent that this should be reconsidered in our approach to paramedic education (Artiga and Hinton 2018; Booske et al. 2010). Additional reasons may include elements of the hidden curriculum that are enacted while undertaking clinical placements. Further exploration of the possible reasons for this trend in our findings is warranted.

Previous literature conducted in other healthcare professions suggests that education, formal training, and clinical exposure are important factors in changing the perceptions of healthcare providers in relation to homelessness (DeLashmutt and Rankin 2005; Hunt 2007; Sibley et al. 2017; Stanley 2013). Clinical experience that involved working with persons experiencing homelessness demonstrated positive changes in the attitudes of nursing students (DeLashmutt and Rankin 2005). The low exposure of participants in our study to homelessness during clinical placement suggests that additional opportunities for interaction with people experiencing homelessness could be considered for integration into the curriculum in order to change perceptions. However, exposure alone may not change perceptions towards homelessness over time (Sibley et al. 2017) - formal education in equality and diversity may also be required to improve knowledge and attitudes when caring for those experiencing homelessness (Jezewski 1995).

There likely exists a need for both formal education and simulated scenarios in paramedic education to facilitate the development of cognitive and affective skills, and the confidence required when caring for this patient population (Leggio and Miller 2017). With appropriate education and experience, paramedics could potentially begin to play a more involved role in the health and social care needs of people experiencing homelessness. For example, they may be well positioned to provide patient education and resources related to access to nutritious meals, personal hygiene issues, healthcare services, and first aid measures. They may be able to provide support in the form of social interventions such as taxi and food vouchers (Bridge 2019). Further insight into the role that paramedics can play in the health and social care of those experiencing homelessness will be best informed through directly involving both of these groups in future research. Paramedics also need to consider their role in the public health and public policy discourse related to social issues such as homelessness. For example, strategic plans to address homelessness or its associated health issues rarely include paramedic services. This is perhaps related to a view of paramedics as emergency responders rather than as healthcare professionals integrated within the health service.

Future studies should aim to enrol larger samples, study practicing paramedics, involve those experiencing homelessness, and consider expanded approaches to data collection. In addition, paramedics need to consider their involvement in policy discussions related to homelessness and broader social issues that have an impact on health. It is our hope that this pilot study encourages an interest in further research in this area of paramedicine.

### Limitations

The use of convenience sampling at a single site, although a simpler recruitment method, means that results may not be representative of paramedic students across our program, or the province. The small sample size means that results may not be generalizable. There are no data on those students who declined to participate. The HPATHI is a self-reported questionnaire that while providing reliable data, does not account for participants’ self-reporting bias. There may be variances in what participants reported, and how they actually conduct themselves in practice. Social desirability bias could have an important effect on results however we attempted to minimize it through an anonymous survey. While the HPATHI was a convenient way to collect our data, the questionnaire itself does not include any statements directly related to paramedic practice. There was a loss of five participants between pre-placement and post-placement questionnaires, and due to our decision to not collect demographic data, we are unable to determine who these participants were.

## Data Availability

Data are available on request.

## Conclusion

The goal of conducting this study was to determine paramedic students’ attitudes towards homelessness in order to suggest improvements in paramedic education and patient care. Our study demonstrated that paramedic students had overall positive attitudes towards people experiencing homelessness, which is encouraging and is the foundation for positive interactions and improved patient outcomes in the future. However, there appeared to be a decreasing trend over time in relation to confidence and interest in working with persons experiencing homelessness. Implementation of curriculum changes could improve paramedic students’ perceptions of caring for patients in this vulnerable population. Further research should be considered into the role paramedics can play in addressing the healthcare and social needs of persons experiencing homelessness.

## Acknowledgements

The authors would like to thank all paramedic students who participated in the study.

## References

Abramovich, A., & Shelton, J. (2017). Where Am I Going to Go? Intersectional Approaches to Ending LGBTQ2S Youth Homelessness in Canada & the U.S. (A. Abramovich & J. Shelton, Eds.). Toronto: Canadian Observatory on Homelessness. http://homelesshub.ca/sites/default/files/Where_Am_I_Going_To_Go.pdf

Artiga, S., & Hinton, E. (2018). Beyond health care: The role of social determinants in promoting health ahd health equity. Kaiser Family Foundation: The Kaiser Commission on Medicaid and the Uninsured, The Essent(May), 1–13. http://files.kff.org/attachment/issue-brief-beyond-health-care

Booske, B. C., Athens, J. K., Kindig, D. A., Park, H., & Remington, P. L. (2010). Country Health Rankings Working Paper: Different perspectives for assigning weights to determinants of health. Population Health Institute, (February).

Bridge, T. (2019, January 31). New cards help paramedics care in other ways. The Stratford Beacon Herald. https://www.stratfordbeaconherald.com/news/local-news/new-cards-help-paramedics-care-in-other-ways

Buck, D. S., Monteiro, F. M., Kneuper, S., Rochon, D., Clark, D. L., Melillo, A., & Volk, R. J. (2005). Design and validation of the Health Professionals’ Attitudes Toward the Homeless Inventory (HPATHI). BMC Medical Education, 5, 1–8. doi:10.1186/1472-6920-5-2

Canadian Observatory on Homelessness. (2019a). Community profiles. Homeless Hub. https://www.homelesshub.ca/CommunityProfiles. Accessed 4 July 2019

Canadian Observatory on Homelessness. (2019b). Youth. About Homelessness. https://www.homelesshub.ca/about-homelessness/population-specific/youth. Accessed 5 July 2019

Canadian Observatory on Homelessness. (2019c). Homelessness defined. Homelessness 101. https://www.homelesshub.ca/about-homelessness/homelessness-101/what-homelessness. Accessed 5 July 2019

City of London. (2017a). Counting our way Home. London, ON. http://s3.documentcloud.org/documents/4173041/170314026-COL-Enumeration-Event-2017-Summary.pdf

City of London. (2017b). London’s Emergency Shelters Progress Report 2011-2016. London, ON. http://s3.documentcloud.org/documents/4173038/Emergency-Shelter-Progress-Report-2017-170823.pdf

City of Toronto. (2018). 2018 Street Needs Assessment. Toronto. https://www.toronto.ca/wp-content/uploads/2018/11/981e-2018-SNA-Results-Highlights-Slides.pdf

Crow, S. (2014). Critical Synthesis Package: Health Professional’s Attitude Towards the Homeless Inventory (HPATHI). MedEdPORTAL Publications. doi:10.15766/mep_2374-8265.9589

DeLashmutt, M. B., & Rankin, E. A. (2005). A different kind of clinical experience: poverty up close and personal. Nurse educator. doi:10.1097/00006223-200507000-00005

Fine, A. G., Zhang, T., & Hwang, S. W. (2013). Attitudes towards homeless people among emergency department teachers and learners: A cross-sectional study of medical students and emergency physicians. BMC Medical Education, 13(1), 1. doi:10.1186/1472-6920-13-112

Frankish, C. J., Hwang, S. W., & Quantz, D. (2005). Homelessness and health in Canada: Research lessons and priorities. Canadian Journal of Public Health, 96(SUPPL. 2). doi:10.2307/41994457

Gaetz, S., Dej, E., Richter, T., & Redman, M. (2016). The state of homelessness in Canada 2016. Toronto. https://homelesshub.ca/sites/default/files/SOHC16_final_20Oct2016.pdf

Goering, P., Tolomiczenko, G., Sheldon, T., Boydell, K., & Wasylenki, D. (2002). Characteristics of Persons Who Are Homeless for the First Time. Psychiatric Services. doi:10.1176/appi.ps.53.11.1472

Hunt, R. (2007). Service-learning: an eye-opening experience that provokes emotion and challenges stereotypes. The Journal of nursing education.

Hwang, S. W., & Bugeja, A. L. (2000). Barriers to appropriate diabetes management among homeless people in Toronto. Cmaj, 163(July 1999), 161–165.

Jezewski, M. A. (1995). Staying Connected: The Core of Facilitating Health Care for Homeless Persons. Public Health Nursing. doi:10.1111/j.1525-1446.1995.tb00010.x

Kimbler, K. J., DeWees, M. A., & Harris, A. N. (2017). Characteristics of the old and homeless: identifying distinct service needs. Aging & Mental Health, 21(2), 190–198. doi:10.1080/13607863.2015.1088512

Leggio, W. J., Giguere, A., Sininger, C., Zlotnicki, N., Walker, S., & Miller, M. G. (2019). Homeless Shelter Users and Their Experiences as EMS Patients: A Qualitative Study. Prehospital Emergency Care, 0(0), 1–6. doi:10.1080/10903127.2019.1626954

Leggio, W. J., & Miller, M. (2017). Homeless Patients and EMS: An Example for Needing Affective Education in EMS. Domain 3, (Summer), 10–12.

Lester, H., & Bradley, C. P. (2001). Barriers to primary healthcare for the homeless: The general practitioner’s perspective. European Journal of General Practice. doi:10.3109/13814780109048777

Marshall, C. (2019). 2019 Homeless Count. Vancouver. https://council.vancouver.ca/20190612/documents/pspc1a-Presentation.pdf

Minick, P., Kee, C. C., Borkat, L., Cain, T., & Oparah-Iwobi, T. (1998). Nurses’ Perceptions of People Who Are Homeless. Western Journal of Nursing Research. doi:10.1177/019394599802000307

Moore, G., Gerdtz, M. F., Hepworth, G., & Manias, E. (2011). Homelessness: Patterns of emergency department use and risk factors for re-presentation. Emergency Medicine Journal, 28(5), 422–427. doi:10.1136/emj.2009.087239

Padgett, D. K., Struening, E. L., Andrews, H., & Pittman, J. (1995). Predictors of emergency room use by homeless adults in New York City: The influence of predisposing, enabling and need factors. Social Science and Medicine. doi:10.1016/0277-9536(94)00364-Y

Parkinson, R. (2009). Nurses’ attitudes towards people who are homeless_: a literature review. Diversity in Health and Care, 6.

Pearson, D. A., Bruggman, A. R., & Haukoos, J. S. (2007). Out-of-Hospital and Emergency Department Utilization by Adult Homeless Patients. Annals of Emergency Medicine, 50(6), 646–652. doi:10.1016/j.annemergmed.2007.07.015

Price, J. H., Desmond, S. M., & Eoff, T. A. (1989). Nurses’ perceptions regarding health care and the poor. Psychological reports, 65(3 Pt 1), 1043–1052.

Redelmeier, D. A., Molin, J.-P., & Tibshirani, R. J. (1995). A randomised trial of compassionate care for the homeless in an emergency department. The Lancet, 345(8958), 1131–1134. doi:10.5555/uri:pii:S0140673695909753

Rich, J. (2017). People experience homelessness, they aren’t defined by it. US Interagency Council on Homelessness. https://www.usich.gov/news/people-experience-homelessness-they-arent-defined-by-it/. Accessed 5 July 2019

Sibley, A., Dong, K. A., & Rowe, B. H. (2017). An Inner City Emergency Medicine Rotation Does Not Improve Attitudes toward the Homeless among Junior Medical Learners. Cureus. doi:10.7759/cureus.1748

Stanley, M. J. (2013). Teaching About Vulnerable Populations: Nursing Students’ Experience in a Homeless Center. Journal of Nursing Education. doi:10.3928/01484834-20130913-03

Tangherlini, N., Villar, J., Brown, J., Rodriguez, R. M., Yeh, C., Friedman, B. T., & Wada, P. (2016). The HOME Team: Evaluating the Effect of an EMS-based Outreach Team to Decrease the Frequency of 911 Use among High Utilizers of EMS. Prehospital and Disaster Medicine. doi:10.1017/S1049023X16000790

The 519. (2019). LGBTQ2S Youth Homelessness in Canada. http://www.the519.org/education-training/lgbtq2s-youth-homelessness-in-canada/in-canada. Accessed 5 July 2019

Wen, C. K., Hudak, P. L., & Hwang, S. W. (2007). Homeless people’s perceptions of welcomeness and unwelcomeness in healthcare encounters. Journal of General Internal Medicine. doi:10.1007/s11606-007-0183-7

Zakrison, T. L., Hamel, P. A., & Hwang, S. W. (2004). Homeless people’s trust and interactions with police and paramedics. Journal of Urban Health, 81(4), 596–605. doi:10.1093/jurban/jth143

Zrinyi, M., & Balogh, Z. (2004). Student nurse attitudes towards homeless clients: A challenge for education and practice. Nursing Ethics. doi:10.1191/0969733004ne707oa

